# Methylenetetrahydrofolate reductase (MTHFR) A1298C polymorphism and risk of lung cancer

**DOI:** 10.1101/19011593

**Authors:** Vandana Rai

## Abstract

Recent epidemiological studies have reported association between methylenetetrahydrofolate reductase (MTHFR) gene polymorphisms and lung cancer. The aim of the present study to perform a meta-analysis of published studies to validate the association between MTHFR A1298C polymorphism and risk of lung cancer.PubMed, Springer Link, Science Direct and Google Scholar databases were searched for eligible studies. Of the 78 initially identified studies, 11 case–control studies with 5,996 patients and 7,404 healthy controls were finally included in the present meta-analysis. Odds ratios (ORs) with 95% confidence intervals (CIs) were estimated to assess the association, and all statistical analyses were performed using MIX software (version 1.7).

No statistically significant associations were found between the MTHFR A1298C polymorphism and lung cancer risk in the additive/ allele contrast, co-dominant/heterozygote, homozygote, dominant and recessive genetic models (C vs. A: OR= 0.95, 95% CI= 0.83-1.08; CC vs. AA: OR= 1.13, 95% CI= 0.83-1.5; AC vs. AA: OR= 0.86, 95% CI= 0.70-1.02; AC+CC vs. AA: OR= 0.89, 95% CI= 0.75-1.05; CC vs. AA+AC: OR= 1.20, 95% CI= 0.89-1.40). Significant heterogeneity between individual studies was evident in all five models. In conclusion, present meta-analysis results indicated that there is no significant association between MTHFR A1298C polymorphism and risk of lung cancer.

## Introduction

Lung cancer is the leading cause of cancer-related death worldwide. The incidence and mortality of lung cancer have been significantly and constantly increasing (Parkin et al.,2002; Jemal et al.,2007; Cui et al.,2011). Lung cancer is still the common cancer in men worldwide (1.1 million cases, 16.5% of the total), with high rates in Central-eastern and Southern Europe, Northern America and Eastern Asia. Very low rates are still estimated in Middle and Western Africa (2.8 and 3.1 per 100,000, respectively)(Ferlay et al.,2010). Lung cancer is a common disease that results from a complex interplay of genetic and environmental risk factors (Kiyohara et al.,2011). Many epidemiological studies have provided evidence that high consumption of vegetables and fruits is associated with a reduced risk of lung cancer (Suzuki et al.,2007). Folate is one of the constituents found in vegetables and fruits, and dietary folate may be one of the micronutrients that provide protection against lung carcinogenesis (Suzuki et al.,2007).

5,10-methyl enetetrahydrofolate reductase (MTHFR) gene (OMIM*607093; chromosome 1p36.3) is an important enzyme involved in folate metabolism and is thought to influence DNA methylation and nucleotide synthesis. The low enzymatic activity of the *MTHFR* C677T genotypic variant is associated with DNA hypomethylation, which may induce genomic instability or randomly reactivates the proto-oncogenes to oncogenes(Ozen et al.,2013). Two common and clinically important polymorphisms (C677T and A1298C) identified in the MTHFR gene (Frosst et al.,1995; Weisberg et al.,1998). Frequency of C677T polymorphism varies greatly worldwide (Rai et al.,2010,2012; Yadav et al.,2017). Substitution at nucleotide 1,298 (exon 7) results in an amino acid substitution of glutamate for alanine at codon 429 (van der Put et al.,1998). A1298C (glutamate to alanine) polymorphism, has been associated with decreased enzyme activity (40%), although to a lesser extent than C677T (Weisberg et al.,1998). A1298C allele frequency differs greatly in various ethnic groups of the world. The prevalence of the A1298C homozygote variant genotype ranges from 7 to 12% in White populations from North America and Europe. Lower frequencies have been reported in Hispanics (4 to 5 %), Chinese (1 to 4 %) and Asian populations (1 to 4%)(Botto and Yang,2000; Robien and Ulrich, 2003). To date, several studies have shown that the MTHFR A1298C polymorphism are associated with either increased or decreased risk of lung cancer, whereas others observed no association between the MTHFR A1298C genotype and lung cancer. Small sample size, various ethnic groups, diet, environment, and methodologies might be responsible for the discrepancy. Therefore, a meta-analysis is required to evaluate MTHFR A1298C polymorphism as risk factor for lung cancer.

## Methods

Present meta-analysis was conducted according to Moose guidelines (Stroup et al.,2000). PubMed, Google Scholar, Springer Link and Elsevier database s were searched for eligible studies. The last search was conducted on December 20, 2016. Following terms were used for search: ‘Methylenetertahydrofolate reductase’, ‘MTHFR’, ‘A1298C’, and ‘lung cancer’.

### Inclusion criteria

The following inclusion criteria were used : (i) study should be case control and should evaluate MTFR A1298C polymorphism, (ii) study should be published, (iii) study should be in English language, (iv) study should contained sufficient data to calculate odds ratio (OR) with 95% confidence interval (CI),and (v)study should not contained duplicated data.

### Data Extraction

The following information was extracted from each included study: first author’s name, journal name, year of publication, country name, number of cases and controls. Number of alleles or genotypes in both cases and controls were extracted or calculated from published data to recalculate ORs.

### Statistical analysis

The associations were indicated as a pooled odds ratio (OR) with the corresponding 95% confidence interval (CI). The heterogeneity between studies was tested using the Q-statistic, which is a weighted sum of the squares of the deviations of individual study OR estimates from the overall estimate (Cochran,1954). When the ORs are homogeneous, Q follows a chi-squared distribution with r – 1 (r is the number of studies) degrees of freedom (df). When P<0.05 then the heterogeneity was considered to be statistically significant. Heterogeneity was quantified with the I^2^ metric (I^2^ = (Q – df)/Q), which is independent of the number of studies in the meta-analysis. I^2^ takes values of between 0 and 100%, with higher values denoting a greater degree of heterogeneity (Zintzaras and Hadjigeorgiou,2005; Zintzaras, 2007). The pooled OR was estimated using fixed effect (FE) (Mantel and Haenszel,1959) and random effect (RE)(DerSimonian and Laird,1986) models. Random effect modeling assumes a genuine diversity in the results of various studies, and it incorporates a between-study variance into the calculations. Hence, when there is heterogeneity between studies then the pooled OR is preferably estimated using the RE model (Whitehead, 2002; Zintzaras, 2007). Genetic models were chosen based on the method described by Thakkinstian et al.(2005), briefly calculating and comparing the ORs of C vs A (allele contrast), CC vs. AA (homozygote), AC vs. AA (co-dominant) and CC+AC vs. AA (dominant) and CC vs. AC+AA (recessive), checking the heterogeneity and significance, then determining the best model (Zhang et al.,2013). The Hardy–Weinberg equilibrium of genotypes of controls was tested and if P >0. 05, then it suggest that the controls followed the Hardy–Weinberg equilibrium (HWE) balance.

### Publication bias

Egger’s test (Egger et al.,1997) and Begg’s test (Begg and Mazumdar,1994) described for funnel plot asymmetry were applied to evaluate the evidence for publication bias. All p values are two tailed with a significance level at 0.05. All statistical analyses were undertaken by MIX version 1.7(Bax et al.,2006).

## Results

### Characteristics of included studies

Information extracted from the studies included in the meta-analysis is provided in tables 1 and 2. Total 78 articles were retrieved using search strategies, but 57 articles did not meet the inclusion criteria after reviewing full articles. Out of remaining twenty one articles, ten studies were also excluded because reported only C677T polymorphism details (Figure 1). Eleven articles were suitable for the inclusion in the meta-analysis (Shen et al.,2001; Siemianowicz et al.,2003; Shen et al.,2005; Shi et al.,2005; Zhang et al.,2005; Hung et al.,2007; Suzuki et al.,2007; Liu et al.,2009; Arslan et al.,2010; Kiyohara et al.,2011; Ozen et al., 2013)(Table 1). Out of eleven studies five studies were from Asian population(Shen et al.,2005; Zhang et al.,2005; Suzuki et al.,2007; Liu et al.,2009; Kiyohara et al.,2011) and remaining studies were from Caucasian population (Shen et al.,2001; Siemianowicz et al.,2003; Shi et al.,2005; Hung et al.,2007; Arslan et al.,2010; Ozen et al., 2013).

**Table 1.** Characteristics of eleven studies included in the present meta-analysis

| Study | Ethnicity | Control | Case | Reference |
| --- | --- | --- | --- | --- |
| Shen et al.,2001 | Non-Hispanic White | 554 | 550 | Can Epidemiol Biomarkers Prev 10:397-401. |
| Siemianowicz et al.,2003 | European | 44 | 146 | Oncol Rep, 10: 1341-4. |
| Shen et al.,2005 | Asian | 49 | 114 | Lung Cancer,49:299-309. |
| Shi et al.,2005 | Non-Hispanic White | 1141 | 1051 | Can Epidemiol Biomarkers Prev 14:1477-84. |
| Zhang et al.,2005 | Asian | 400 | 505 | Acta Acad Med Sin 27:700-703. |
| Hung et al.,2007 | European | 2865 | 2209 | Carcinogenesis 28:1334–1340 |
| Suzuki et al.,2007 | Asian | 1019 | 485 | Carcinogenesis 28:1718–1725. |
| Liu et al.,2009 | Asian | 716 | 358 | Can. Geno & Proteo 6: 325-330. |
| Arslan et al.,2011 | European | 61 | 64 | Mol Biol Rep (2011) 38:991–996 |
| Kiyohara et al.,2011 | Asian | 379 | 462 | BMC Cancer 2011, 11:459 |
| Ozen et al.,2013 | European | 176 | 52 | Asian Pac J Cancer Prev, 14 (9):5449-5454 |

**Table 2.**
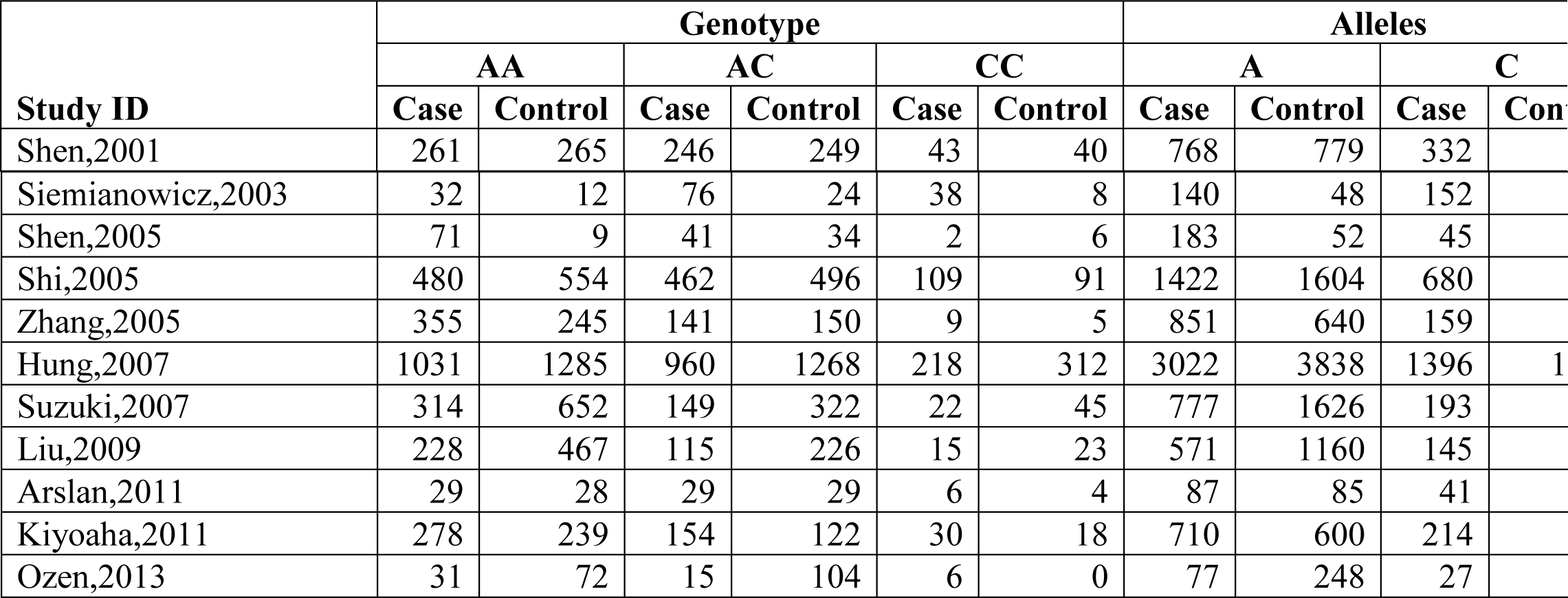
The distributions of MTHFR A1298C genotypes and alleles number for lung cancer cases and controls

**Figure 1.**
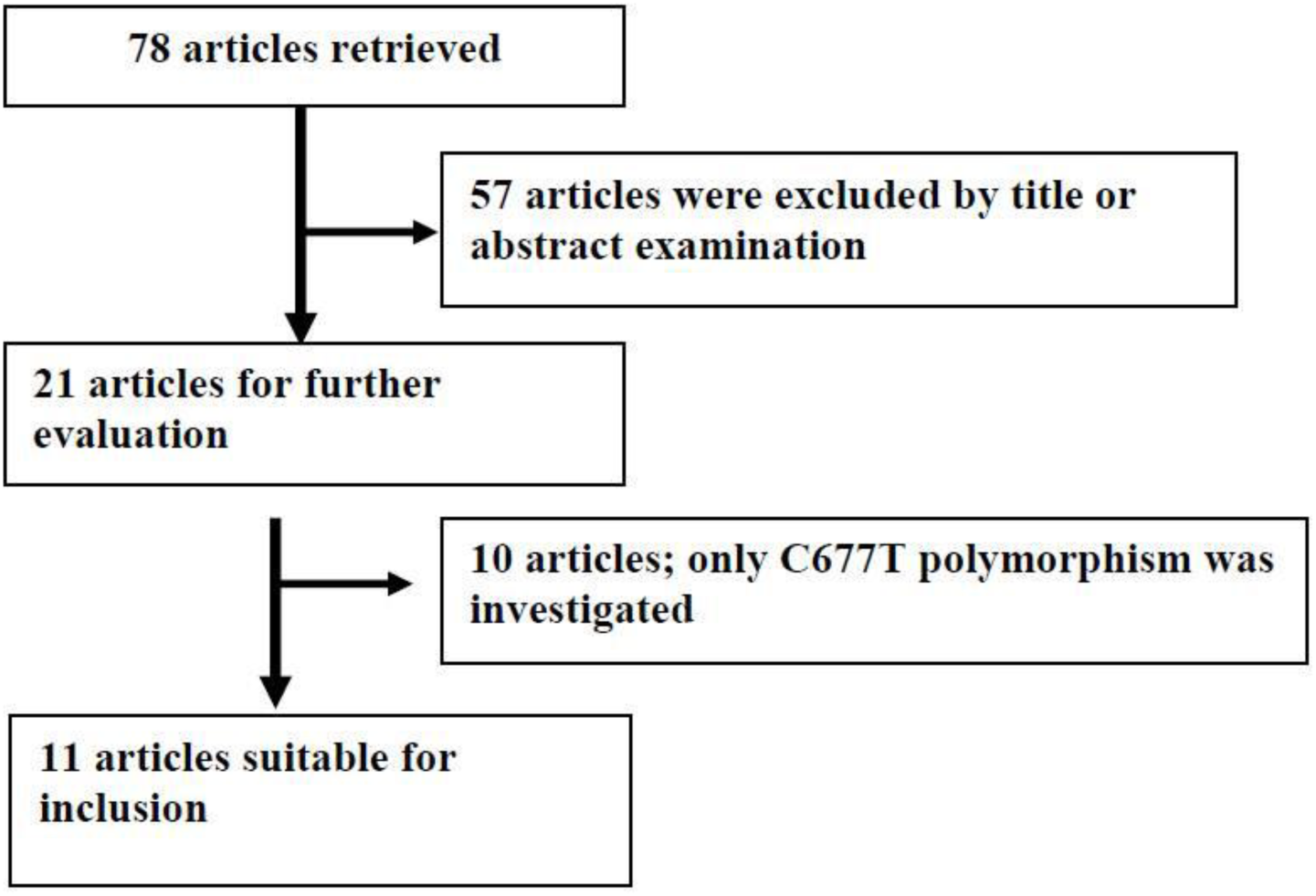
Flow diagram of study selection.

Overall, eleven studies provided 5,996/7404 cases/controls for MTHFR A1298C polymorphism with AA (3,110), AC (2,388) and CC (498) genotypes in cases, and with AA (3,828), AC (3,024), and CC (552) genotypes in controls. In total cases, genotype percentage of AA, AC, and CC was 51.67%, 38.83% and 8.30% respectively. In controls genotypes, percentage of AA, AC and CC were 51.70%, 40.84%, and 7.45% respectively. The frequencies of the genotypes AA and AC were the highest in both cases and controls, and allele A was the most common (Table 2). In all the studies, distribution of genotypes in the control group was in Hardy Weinberg Equilibrium.

### Meta-analysis

Meta-analysis with allele contrast (C vs A) showed no significant association with both fixed effect (OR _C vs A_= 0.99; 95%CI= 0.93-1.04; p= 0.062; P_Pb_= 0.44) and random effect model (OR _C vs A_ = 0.95; 95% CI= 0.83-1.08; p= 0.44) (Table 3, Figure 2).

**Table 3:** Summary estimates for the odds ratio (OR) of MTHFR A1298C in various allele/genotype contrasts, 95% confidence limits, the significance level, p value of heterogeneity test (Q test), and the I^2^ metric, and publication bias p-value (Egger Test).

| Genetic Models | Fixed effect<br>OR (95% CI), p<br>value | Random effect<br>OR (95% CI),p<br>value | Heterogeneity p-value<br>(Q<br>test) | $I^2$<br>(%) | Publication bias<br>(p value of<br>Begg's<br>test) | Publication bias (p<br>of Egger's<br>test) |
| --- | --- | --- | --- | --- | --- | --- |
| Additive (C vs A) | 0.99(0.93-1.04),0.62 | 0.95(0.83-1.08),0.44 | <0.0001 | 75.12 | 0.24 | 0.44 |
| Co-dominant (AC vs AA) | 0.94(0.87-1.0),0.07 | 0.86(0.7-1.02),0.9 | <0.0001 | 73.77 | 0.07 | 0.13 |
| Homozygote (CC vs AA) | 1.05(0.92-1.2),0.5 | 1.13(0.83-1.5),0.42 | 0.002 | 64.03 | 0.75 | 0.51 |
| Dominant (CC+AC vs AA) | 0.95(0.88-1.01),0.16 | 0.89(0.75-1.05),0.19 | <0.001 | 75.05 | 0.19 | 0.23 |
| Recessive (AA+AC vs CC) | 1.1(0.93-1.2),0.35 | 1.2(0.89-1.4),0.27 | 0.02 | 53.36 | 0.63 | 0.25 |

**Figure 2.**
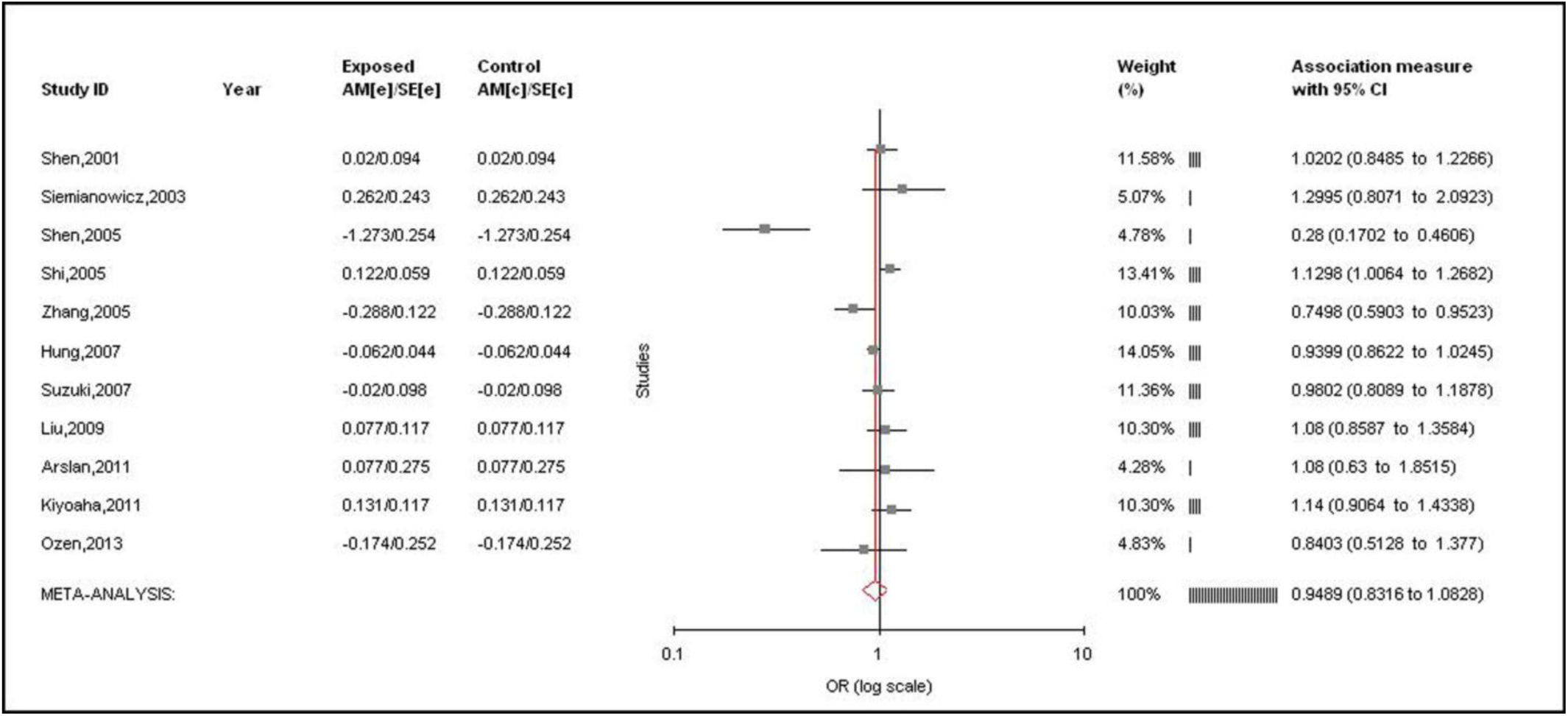
Forest plots for the association between MTHFR A1298C polymorphism and lung cancer for allele contrast model (C vs A) with random effect model.

Table 3 summarizes the ORs with corresponding 95% CIs for association between mutant A1298C polymorphism and risk of lung cancer in homozygote, co-dominant, dominant, and recessive models. Genotype meta-analysis did not report any association with lung cancer (CC vs AA (homozygote model): OR_CC vs AA_ = 1.13, 95%CI= 0.83-1.5, p= 0.42 (Figure 3); AC vs. AA (co-dominant model): OR_AC vs AA_ = 0.86, 95%CI= 0.70-1.02, p= 0.90; CC+AC vs. AA (dominant model): OR_CC+AC vs AA_ = 0.89, 95%CI= 0.75-1.05, p= 0.19; CC vs AC+AA (recessive model): OR_CC vs AC+AA_ = 1.2, 95%CI= 0.89-1.4, p= 0.27 (Figure 4)).

**Figure 3.**
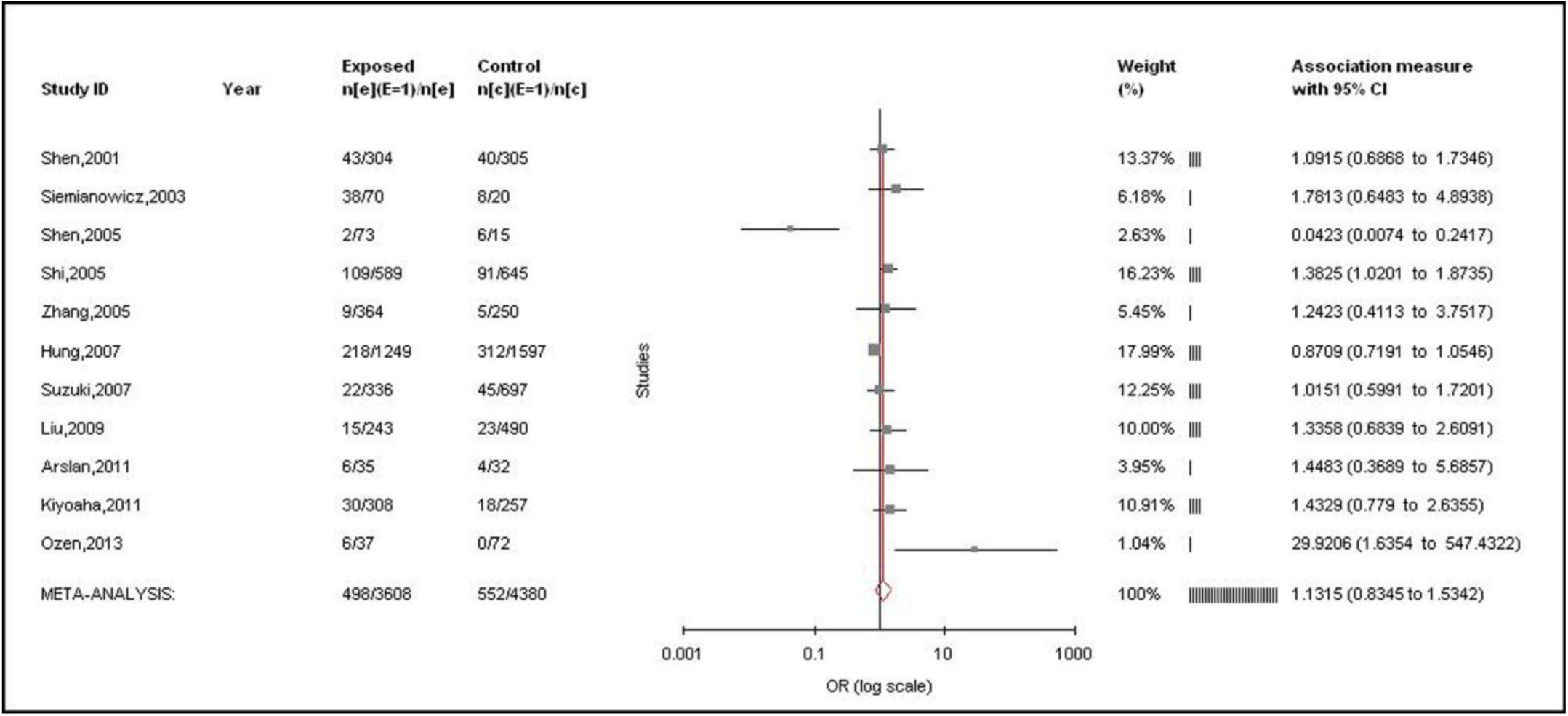
Forest plots for the association between MTHFR A1298C polymorphism and lung cancer for homozygote model (CC vs AA) with random effect model.

**Figure 4.**
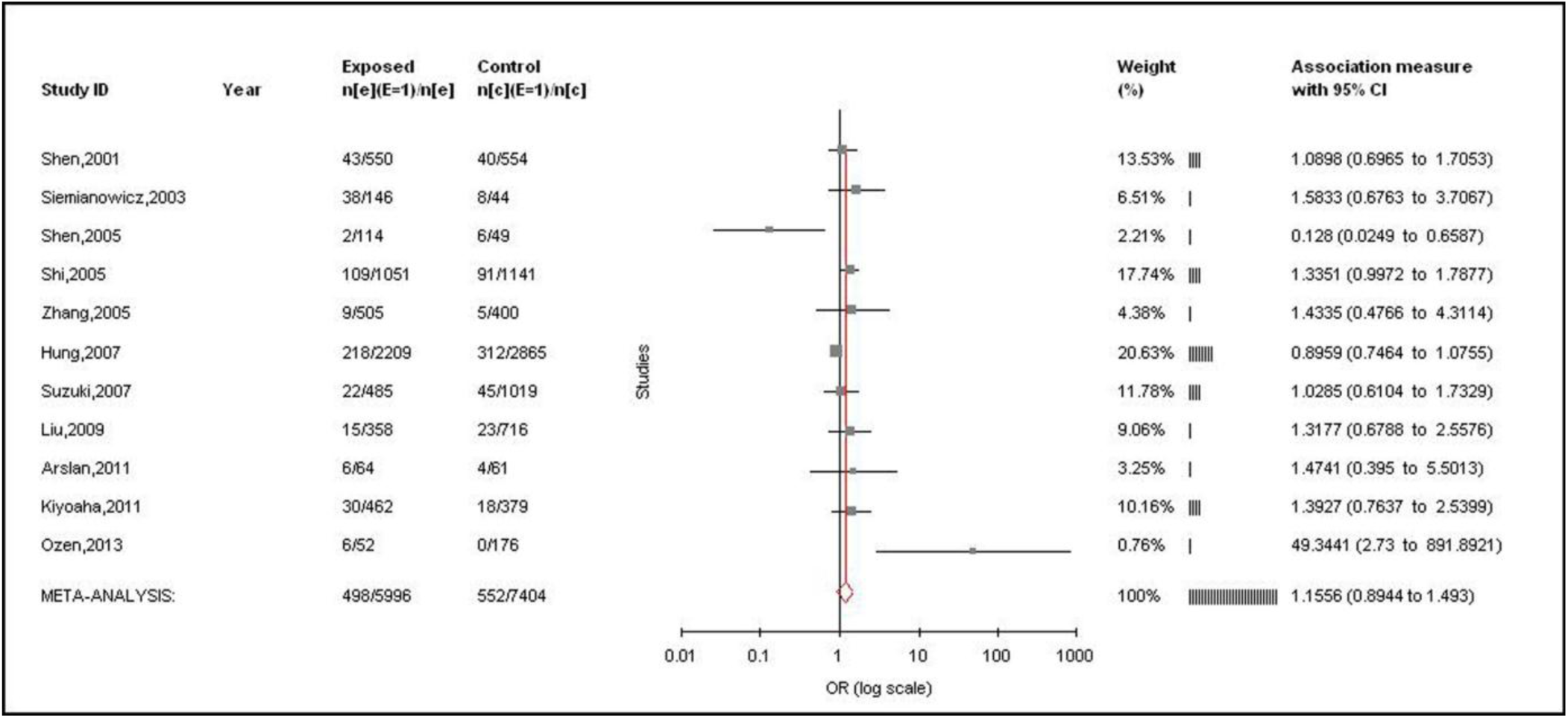
Forest plots for the association between MTHFR A1298C polymorphism and lung cancer for dominant model (CC +AC vs AA) with random effect model.

A true heterogeneity existed between studies for allele contrast (P_heterogeneity_ <0.0001, Q= 40.19, I^2^=67.22%, t^2^=0.030, z =0.78) and genotype homozygote (P_heterogeneity_ =0.002, Q= 27.83, I^2^=64.07%, t^2^=0.12, z = 0.79), dominant (P_heterogeneity_ <0.0001, Q= 40.08, I^2^= 75.05%, t^2^=0.05, z = 1.30) and recessive (P_heterogeneity_ =0.02, Q= 21.44, I^2^= 53.36%, t^2^=0.07, z = 1.1) comparisons. The ‘I^2^’ value of more than 50% for between studies comparison in both allele and genotype analysis shows high level of true heterogeneity.

### Publication bias

Funnel plots using standard error and precision values for allele and genotypes using random effect model were generated (Figure 5). Symmetrical distribution of studies in the funnel plots suggests absence of publication bias. This is also supported by Beggs and Eggers test (Begg’s p= 0.24, Egger’s p= 0.44 for C vs. A; Begg’s p= 0.75, Egger’s p= 0.51 for CC vs AA; and Begg’s p= 0.07, Egger’s p= 0.13 for AC vs. AA; Begg’s p= 0.19, Egger’s p= 0.23 for CC+AC vs. AA; Begg’s p= 0.63, Egger’s p= 0.25 for CC vs. AC+AA) (Table 3).

**Figure 5.**
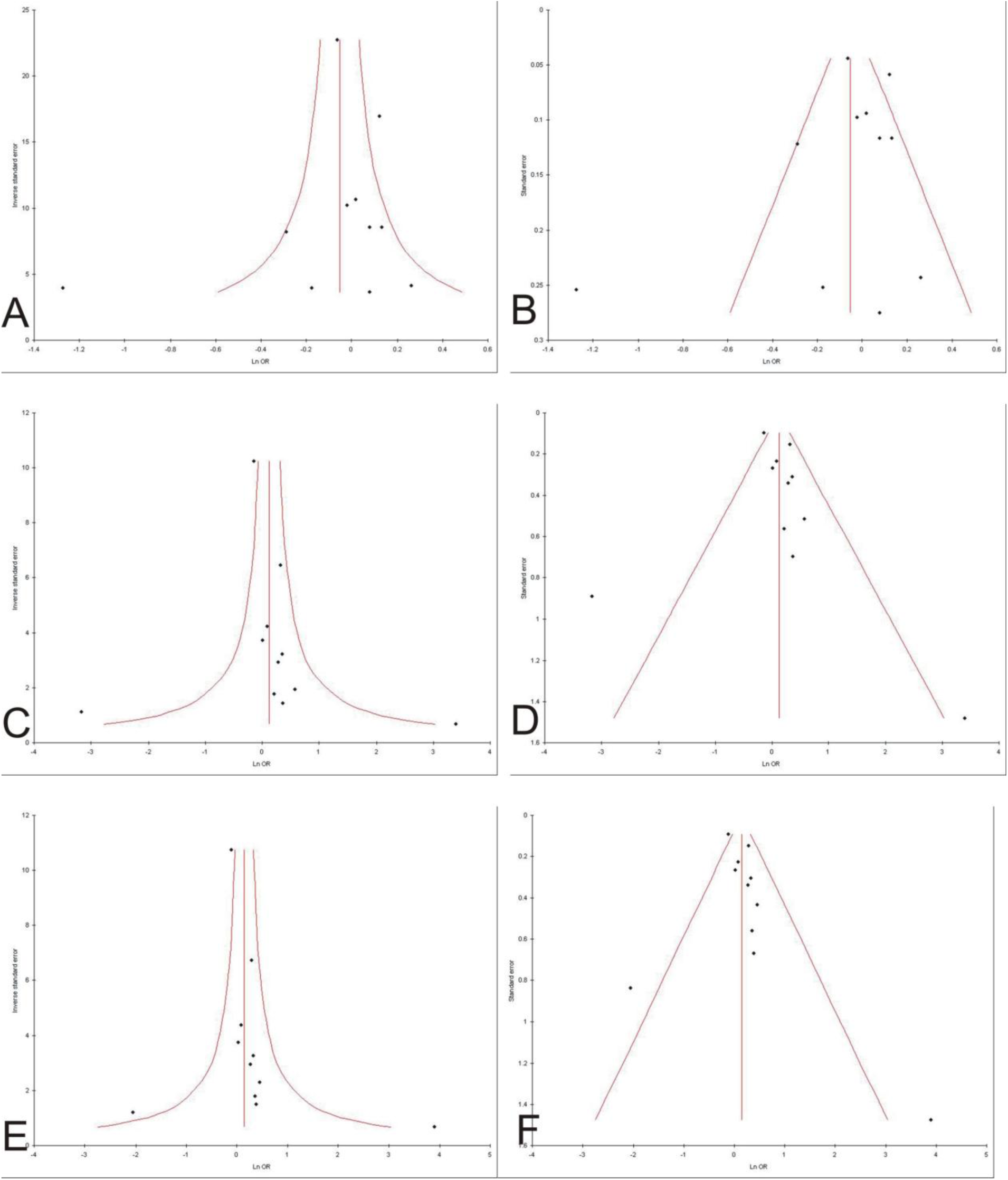
Funnel plots A. precision versus OR (C vs A), B. standard error versus OR (C vs A) C. precision versus OR(CC vs AA), D. standard error versus OR (CC vs AA), E. precision versus OR (CC+AC vs AA), F. standard error versus OR (CC+AC vs AA).

## Discussion

MTHFR plays a central role in balancing DNA synthesis (which involves 5,10-methylentetrahydrofolate) and DNA methylation (which involves 5,10-methyltetrahydrofolate). Specifically, the 677T allele contributes to DNA hypomethylation, which in turn may lead to altered gene expression; at the same time, this polymorphism might exert a protective effect, as observed for colorectal cancer (Botto and Yang, 2000), by increasing the levels of the MTHFR substrate, essential for DNA synthesis. Folate deficiency and metabolism disorders may cause DNA hypomethylation, and A to C substitution at nucleotide 1298 in MTHFR, which alters enzyme activity, affecting DNA methylation or DNA synthesis, thereby increasing susceptibility to cancer (Umar et al.,2010; Ekiz et al.,2012; Tan et al.,2013). Present meta-analysis included eleven studies with a total of 5,996 cases and 7,404 controls have investigated the association between A1298C polymorphism with lung cancer.

Meta-analysis is a powerful tool for analyzing cumulative data of studies wherein the individual sample sizes are small and the disease can be easily masked by other genetic and environmental factors (Liang et al.,2013). A meta-analysis potentially investigates a large number of individuals and can estimate the effect of a genetic factor on the risk of the disease (Liang et al., 2013). Several meat-analyses were published to asses effect of folate pathway genes polymorphism as risk for several diseases like-MTHFR prevalence (Yadav et al., 2017),breast cancer (Rai,2014; Kumar et al., 2015; Rai et al., 2017), Ovary cancer (Rai,2016), prostate cancer (Yadav et al.,2016), colorectal cancer (Rai,2015), glucose-6 phosphate dehydrogenase deficiency (Kumar et al.,20016), recurrent pregnancy loss (Rai,2016), hyperurecemia (Rai,2016), Down syndrome (Rai,2011; Rai et al.,2017; Rai and Kumar, 2018), cleft lip/palate (Rai,2014,2017), NTD (Yadav et al., 2015), epilepsy (Rai and Kumar, 2018), schizophrenia (Yadav et al.,2016; Rai et al.,2017), autism (Rai, 2016; Rai and Kumar,2018), depression (Rai,2014), Alzheimers disease (Rai,2016), male infertility (Rai and Kumar,2017), prostate cancer (Yadav et al., 2016), uterine leiomyioma (Kumar and Rai,2018),digestive tract cancer (Yadav et al., 2018), endometrial cancer (Kumar et al., 2018) and esophageal cancer (Kumar and Rai,2018) etc.

### Limitations

(i) sample size in few studies were small, (ii)controls were not uniform in all studies, in some studies hospital based patients of other diseases were considered, (iii) other important factors like smoking and folate intake were not considered in the present meta-analysis and (iv) present review is restricted only one folate pathway gene polymorphism. Further the main strength of the present meta-analysis is absence of publication bias and larger pooled sample size. Present meta-analysis suggested that A1298C polymorphism did not play any role in the etiology of lung cancer.

## Data Availability

All data included in the manuscript

## Conflict of Interest

**None**

## Notes

### Competing Interest Statement

The authors have declared no competing interest.

### Funding Statement

No Funding

